# Health Education for Parents During the COVID-19 Outbreak Public Health Education for Parents During the Outbreak of COVID-19: A Rapid Review

**DOI:** 10.1101/2020.04.14.20064741

**Authors:** Weiguo Li, Jing Liao, Qinyuan Li, Muna Baskota, Xingmei Wang, Yuyi Tang, Qi Zhou, Xiaoqing Wang, Xufei Luo, Yanfang Ma, Toshio Fukuoka, Hyeong Sik Ahn, Myeong Soo Lee, Yaolong Chen, Zhengxiu Luo, Enmei Liu, on behalf of COVID-19 evidence and recommendations working group

**Affiliations:** Department of Respiratory Medicine, Children’s Hospital of Chongqing Medical University, Chongqing 400014, China; National Clinical Research Center for Child Health and Diseases, Ministry of Education Key Laboratory of Child Development and Disorders, China International Science and Technology Cooperation Base of Child Development and Critical Disorders, Children’s Hospital of Chongqing Medical University, Chongqing 400014, China; Chongqing Key Laboratory of Pediatrics, Chongqing 400014, China; The First Clinical Medical College of Lanzhou University, Lanzhou, China; Evidence-based Medicine Center, School of Basic Medical Sciences, Lanzhou University, Lanzhou 730000, China; School of Public Health, Lanzhou University, Lanzhou 730000, China; Emergency and Critical Care Center, the Department of General Medicine, Department of Research and Medical Education at Kurashiki Central Hospital, Japan; Advisory Committee in Cochrane Japan, Japan; Department of Preventive Medicine, Korea University College of Medicine, Seoul, Korea; Korea Cochrane Centre, Korea; Korea Institute of Oriental Medicine, Daejeon, Korea; University of Science and Technology, Daejeon, Korea; Lanzhou University, an Affiliate of the Cochrane China Network, Lanzhou 730000, China; Chinese GRADE Center, Lanzhou 730000, China; Key Laboratory of Evidence Based Medicine and Knowledge Translation of Gansu Province, Lanzhou University, Lanzhou 730000, China

**Keywords:** COVID-19, children, health education, parents, rapid review

## Abstract

**Background:** It is well-known that public health education plays a crucial role in the prevention and control of emerging infectious diseases, but how health providers should advise families and parents to obtain health education information is a challenging question. With COVID-19 (Coronavirus disease 2019) spreading around the world, this rapid review aims to answer that question and thus to promote evidence-based decision making in health education policy and practice.

**Methods:** We systematically searched the literature on health education during COVID-19, SARS (severe acute respiratory syndrome) and MERS (middle east respiratory syndrome) epidemics in Medline (via PubMed), Cochrane Library, EMBASE, Web of Science, CBM (China Biology Medicine disc), CNKI (China National Knowledge Infrastructure), and Wanfang Data from their inception until March 31, 2020. The potential bias of the studies was assessed by Joanna Briggs Institute Prevalence Critical Appraisal Tool.

**Results:** Of 1067 papers found, 24 cross-sectional studies with a total of 35,967 participants were included in this review. The general public lacked good knowledge of SARS and MERS at the early stage of epidemics. Some people’s knowledge, attitude and practice (KAP) of COVID-19 had been improved, but the health behaviors of some special groups including children and their parents need to be strengthened. Negative emotions including fear and stigmatization occurred during the outbreaks. Reliable health information was needed to improve public awareness and mental health for infectious diseases. Health information from nonprofit, government and academic websites was more accurate than privately owned commercial websites and media websites.

**Conclusions:** For educating and cultivating children, parents should obtain information from the official websites of authorities such as the World Health Organization (WHO) and national Centers for Disease Control, or from other sources endorsed by these authorities, rather than from a general search of the internet or social media.

## Background

Since late 2019, the COVID-19 outbreak has become the most urgent public health issue threatening lives over the world (1). The disease is caused by SARS-CoV-2, a new type of coronavirus, and it usually presents manifestations of pneumonia that takes a severe form in approximately 20 percent of confirmed patients (2). As of March 30th, 2020, over 720,000 laboratory-confirmed COVID-19 patients have been diagnosed globally, and the number will probably continue to increase until the epidemic trend tempers, which caused a significant impact on the health care and economy of affected areas. With over 200 countries affected and thousands of deaths, World Health Organization (WHO) declared the outbreak as a global pandemic on March 11, 2020 (3). It is well-known that public health education plays a crucial role in the prevention and control of infectious diseases, but how health providers advise families or parents to obtain health education information is a challenging question. In the context of a public health emergency, health education practice was often neglected or unprepared. This rapid review focuses on this topic to answer that question and aims to promote evidence-based decision making in health education policy and practice.

A rapid review has emerged as a helpful approach to provide actionable and relevant evidence in a timely and cost-effective manner (4). Rapid reviews, as well as rapid advice guidelines, should be conducted in a limited time to offer prompt evidence for the public health emergency. This review intends to support a rapid advice guideline for children with COVID-19 (5) about how health providers should advise families and parents to obtain health education information on SARS-CoV-2 infection. There were various existing methods to expedite the conduct of a rapid review, yet we followed the WHO rapid review practical guide (4).

## Methods

The pre-search found that the most original studies on the topic were cross-sectional surveys, and the participants were people from non-specific groups rather than parents. However, a health education strategy is urgently needed to help prevent the disease. So, it was implied that this rapid review would use indirect evidence to draw the conclusion. We expanded the search range to SARS and MERS, since these two other coronaviruses have similar transmission routes and clinical outcomes as SARS-CoV-2.

### Search strategy

Three researchers (W Li, J Liao and Q Li) independently performed a comprehensive search of the following electronic databases: Medline (via PubMed), Cochrane Library, EMBASE, Web of Science, CBM (China Biology Medicine disc), CNKI (China National Knowledge Infrastructure), and Wanfang Data from their inception until March 31, 2020. We only included studies published in Chinese or English. Using the Boolean approach, the databases will be searched for the following key terms in titles or abstracts: (Health Education OR Health Promotion OR Parent Education) AND (2019-novel coronavirus OR 2019-ncov OR COVID-19 OR SARS* OR MERS* OR Severe Acute Respiratory Syndrome OR Middle East Respiratory Syndrome). We also searched Google Scholar and reference lists of relevant reviews for unpublished or further potential studies (*The details of the search strategy can be found in the Supplementary Material 1)*.

### Eligibility criteria

We included observational studies, such as cohort and cross-sectional studies, that focused on the health education and health promotion issues during COVID-19, SARS, and MERS epidemics. We included studies with participants from the general public. We excluded studies only focusing on specific groups, like different occupations. Duplicates, studies with specific data missing, studies where full-text was unavailable, comments and letters were also excluded.

### Study selection

After eliminating duplicates, two reviewers (W Li and J Liao) independently performed the search in two steps. Any discrepancies were discussed or solved with a third researcher (Q Li). We used the bibliographic software Endnote. In Step 1, all titles and abstracts were screened using the pre-defined criteria, and studies will be sub-categorized into three (potentially eligible, excluded, and unsure) groups. In Step 2, full-texts of potentially eligible and unclear studies were reviewed to identify the final inclusion. All reasons for exclusion of ineligible studies were recorded, and the process of study selection was documented using a PRISMA flow diagram (6).

### Data extraction

Two researchers extracted the data independently using a structured form. Then the forms were cross-checked to ensure the data accuracy. If any disagreements occurred, they were resolved by discussion. We extracted the following data: 1) study characteristics, including the first author, publication year, and the country or area; 2) participants’ characteristics; 3) aims of survey, and the sample size; and 4) the education resources mentioned in surveys.

### Risk of bias assessment

Two researchers (W Li and J Liao) independently assessed the potential bias of the studies. All discrepancies in quality ratings were discussed with a third researcher (Q Li). Based on the results of the pre-test, we used the Joanna Briggs Institute Prevalence Critical Appraisal Tool (7), which is specifically used for systematic reviews of prevalence studies. Sample representativeness, sampling method, sample size, study subjects and setting, coverage of the identified sample, using of standardized criteria for the measurement, reliability of the measurement, appropriateness of the statistical analysis, confounding factors/subgroups/differences, and objectiveness of the criteria of subpopulations were evaluated in all included studies.

### Data synthesis

For dichotomous outcomes we calculated the risk ratio (RR) and the corresponding 95% confidence interval (CI) and *P* value. For continuous outcomes, we calculated the mean difference (MD) and its corresponding 95% CI when means and standard deviations (SD) were reported. If sufficient data were available, we considered examined the robustness of meta-analyses in a sensitivity analysis.When effect sizes could not be pooled, we reported the study effectives narratively.

### Quality of the evidence assessment

The quality of evidence for each outcome was assessed using the GRADE (The Grading of Recommendations Assessment, Development and Evaluation) approach (8). The criteria mainly considered included study methodological quality, directness of the evidence, heterogeneity of data, precision of effect estimates, and risk of publication bias. The quality of evidence for each outcome was graded as high, moderate, low, or very low.

As COVID-19 is a public health emergency of international concern and the situation is evolving rapidly, our study was not registered in order to speed up the process.

### Study results

Our rapid review included 24 cross-sectional survey studies (9-32) (*Figure 1 and Table 1*) with a total of 35,967 participants, conducted during the COVID-19, SARS and MERS epidemic. Six anonymous network sampling surveys (9-14) were conducted during the COVID-19 outbreak, and the other 18 surveys (15-32) were for SARS and MERS. Nine studies (11,13,15,16,18,20,22,43,44) mentioned the channels from where people obtained health education information. We also found out three studies about health education campaigns (33-35).

**Table 1.**
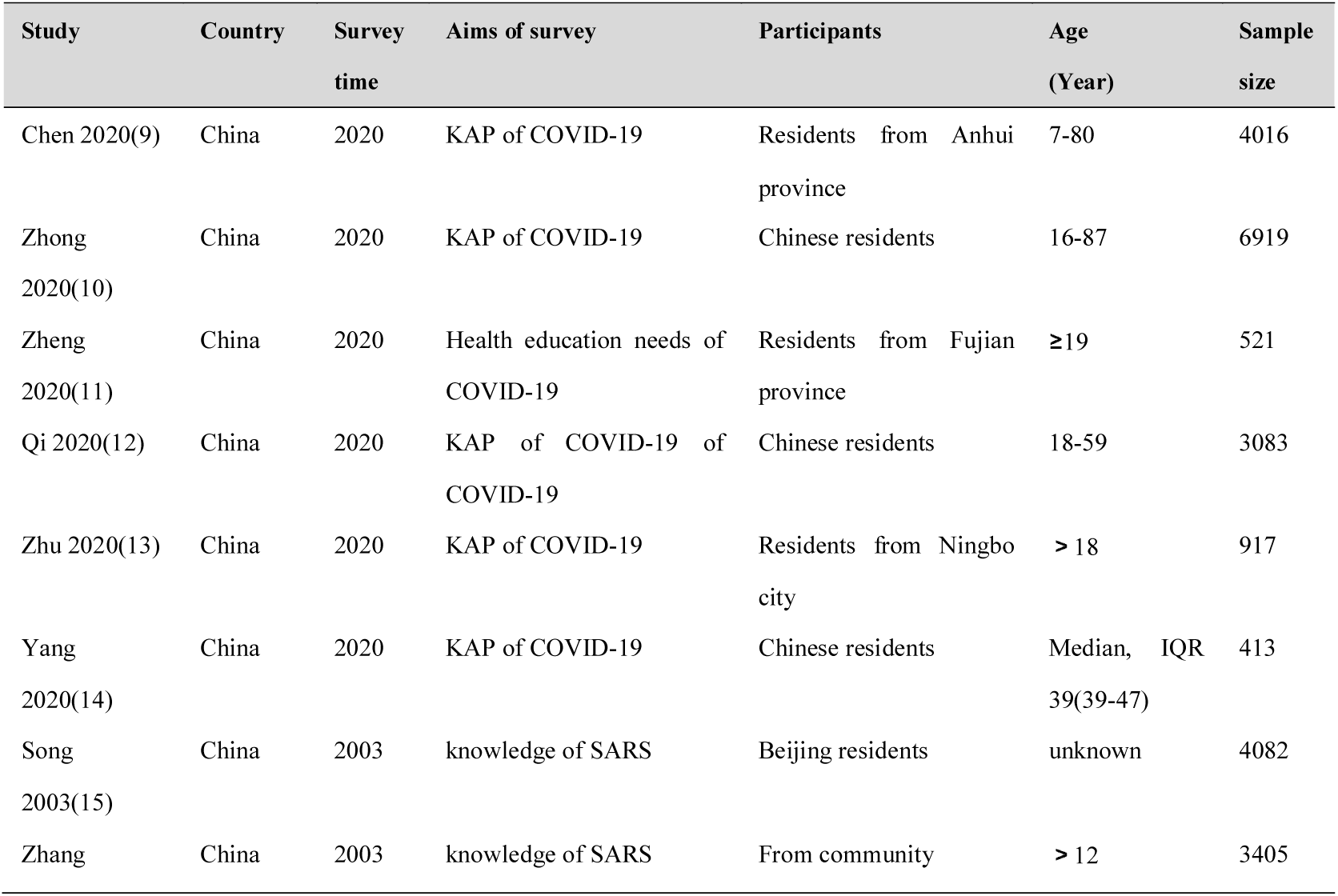
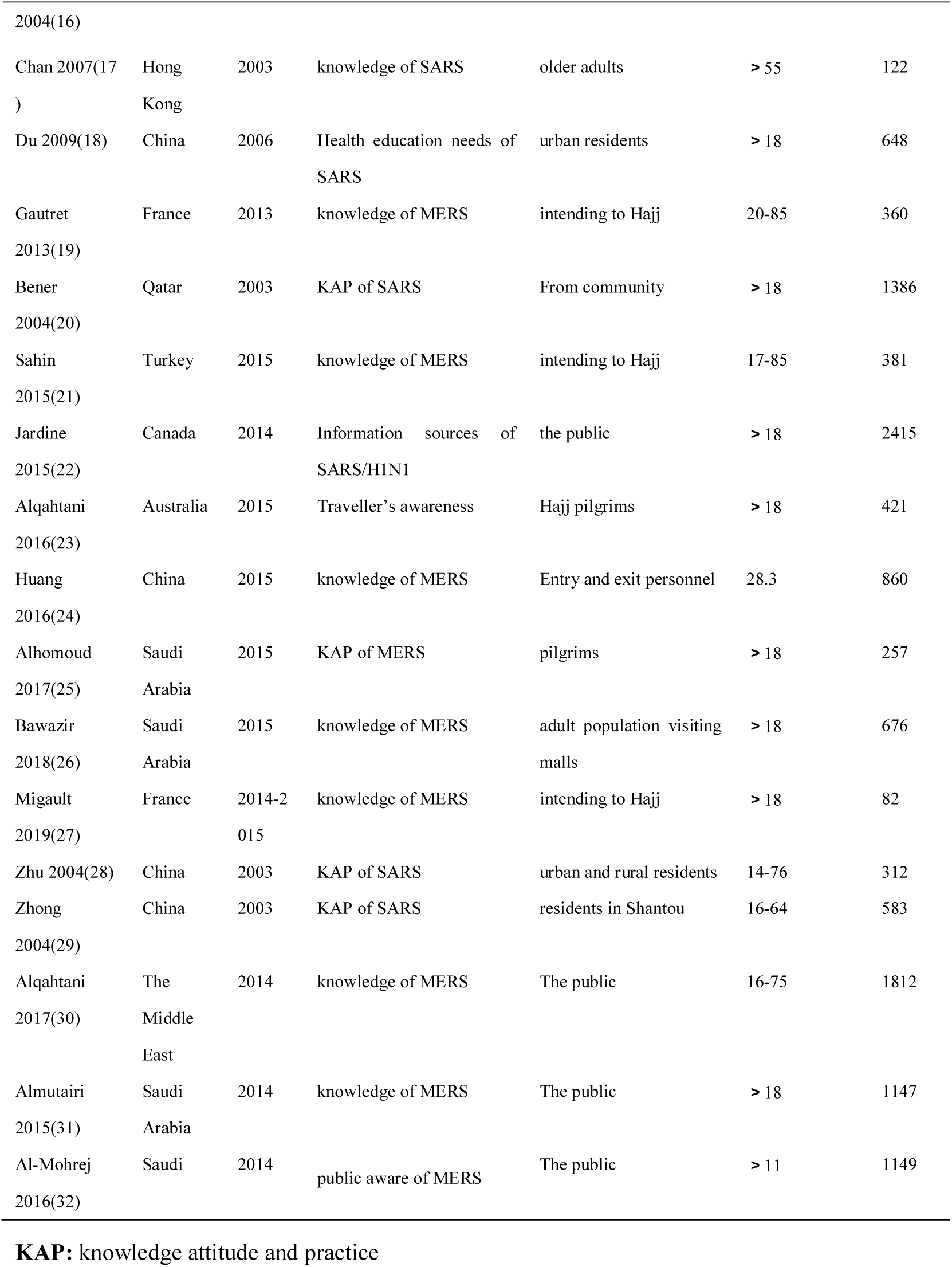
Basic Characteristic of Cross-sectional surveys for awareness and knowledge on COVID-19, SARS and MERS.

**Figure 1.**
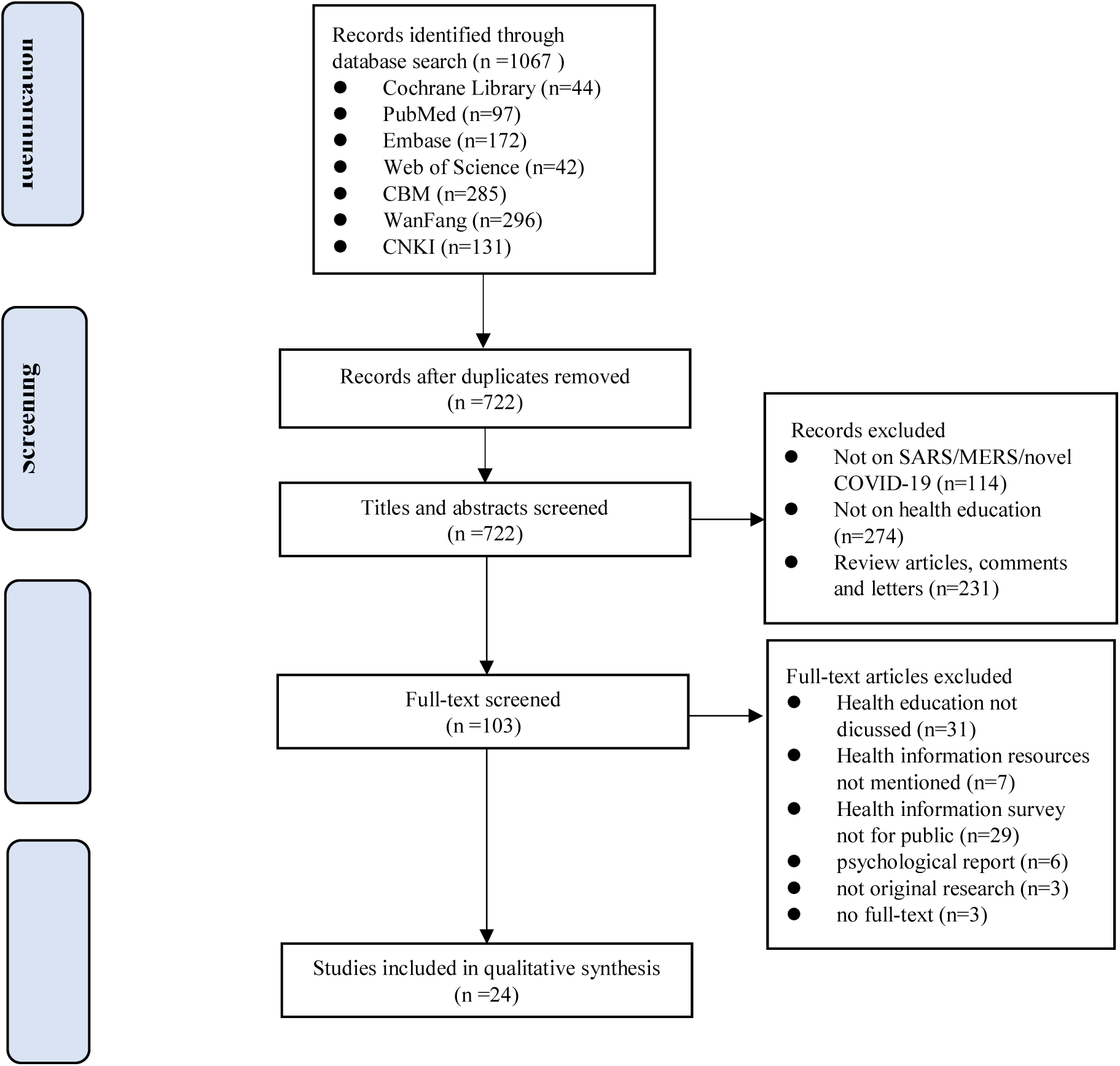
Flow diagram for study selection process

### The public lack of awareness of infectious diseases

Seven studies (15,20,24,26,30-32) revealed that the public did not know how to face emerging infectious diseases. However, three surveys (9,10,12) showed that people had good knowledge attitude and practice (KAP) of COVID-19, yet it is necessary to strengthen the community publicity, and the health education of residents. One survey (10) revealed that most Chinese residents are knowledgeable about COVID-19, hold optimistic attitudes, and have appropriate practices (like wearing mask) towards COVID-19. One study (36) showed that myths, fear and stigmatization of potential SARS patients emerged in the outbreak, as global media reported dramatic stories from the media, television, and the Internet. Another study (37) revealed that emerging health hazards were over-reported in mass media in comparison to the common threats to public health. A survey (26) from Saudi Arabia suggested that epidemiological knowledge received by the public was inadequate, and their recognition of MERS-CoV relied on the clinical manifestations, rather than epidemic features. Age≤30 years, male gender, and not having tertiary education were independent predictors of poor epidemiological knowledge.

Five studies (19,21,23,25,27) showed that pilgrimage travelers lacked awareness of infectious diseases. The Middle East, where MERS outbroke, is also a region with a great number of religious pilgrimage sites. A study in Australia evaluated pilgrims’ awareness of MERS-CoV during travel, preventive measures, and the contact with camel exposure, and found that only 28.0% of Australian pilgrims knew that Saudi Arabia was affected by the MERS-CoV outbreak (23). A survey in France (19) reported that only 35.3% of people knew about the Saudi Ministry of Health recommendations for at-risk pilgrims to postpone participation in the 2013 Hajj. None of the 179 at-risk individuals decided to cancel their Hajj participation even after being advised during the consultation. A study from the United States showed that their CDC’s health website for visitors was frequently clicked in the first half of 2003, and visitors visited 2.6 million times for travel warnings, consultations, and other SARS-related documents (41).

### The communication mode and reliability of health education

We explored the current information-seeking strategies and preferences. Two surveys (11,13) showed that participants tended to obtain the COVID-19 information through Internet, social media by cellphone. One survey (18) from China showed that the top five main channels for the public to obtain information during the SARS period were television (85.4%), interpersonal communication (53.4%), newspapers (48.5%), radio (39.6%) and conference communication (22.6%). People generally preferred the three traditional major types of media to acquire knowledge of health education when SARS outbreak. Another study from China (16) investigated the KAP of SARS prevention and control in the late period of the SARS epidemic and found that people acquired SARS-related knowledge through a variety of media, including the Internet, forums, telephone, television, and newspapers. Voeten *et a*l (42) studied the sources of information, epidemiological knowledge, and health concepts of Chinese in Europe during epidemics of infectious diseases. The study found that in the case of SARS and bird flu outbreaks in China, people acquired most of the information from their relatives and friends, followed by local unofficial networks and Chinese television stations in the United Kingdom and the Netherlands. Another telephone survey from Canada (22) compared the sources of information used by the public during the SARS epidemic in 2003 and the H1N1 epidemic in 2010 and found that traditional mass media were still the most commonly used sources for information in both groups of surveys. Information could be also obtained from friends and relatives, but they were not considered very useful or credible. More and more people are using multiple sources for information: the proportion of people following at least five sources for information has increased by 60.0% between 2003 and 2010.

### The effect of health education

A survey (10) showed health education programs aimed at improving COVID-19 knowledge are helpful for Chinese residents to hold optimistic attitudes and maintain appropriate practices. During the SARS epidemic in 2003, a study evaluated the level of the awareness of SARS among older people, then provided them with health education by telephone, and assessed the change. The authors found that after telephone health education, the anxiety level of the elderly people decreased, and their awareness of the modes of virus transmission improved (17). A study from China showed that comprehensive health education and publicity improved Beijing residents’ attitude towards SARS, enhanced their awareness of preventing SARS, and significantly decreased the occurrence rates of tension, anxiety, depression and fear (*P*<0.01). There were also more behavior changes (*P*<0.01) in the prevention of SARS, which fully testifies the effect of comprehensive health education publicity (15). Another study showed that the awareness of four items of MERS knowledge was significantly higher after a health education intervention than before (*P*<0.01). The health behavior formation rate of MERS was also higher after the intervention than that before (*P*<0.01) (23).

### Health education and health promotion programs launched after SARS in 2003

The world has experienced several acute public health emergencies requiring global coordination. Since the global epidemic of SARS in 2003, many institutions and organizations have launched health education programs (*Table 2*). It is worth mentioning that the British government launched a public information campaign on coronavirus quickly after the diagnosis of the first local SARS-CoV-2 infection to inform citizens how to slow the spread of SARS-CoV-2 in the UK.

**Table 2.**
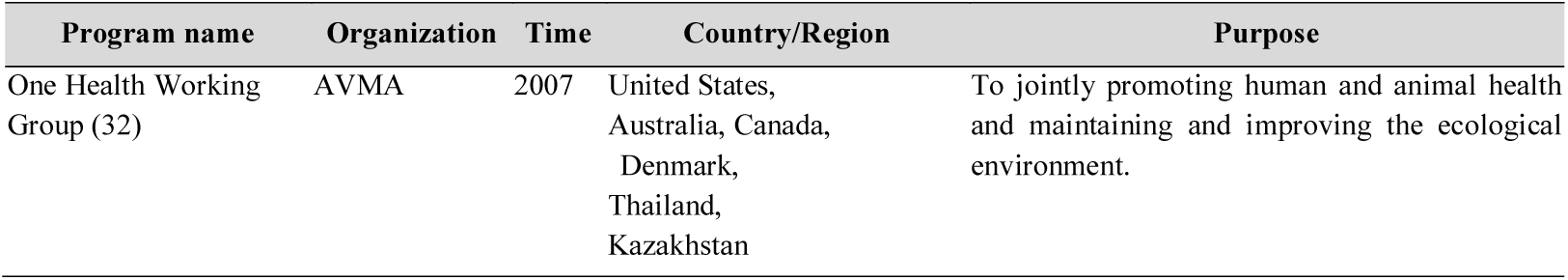
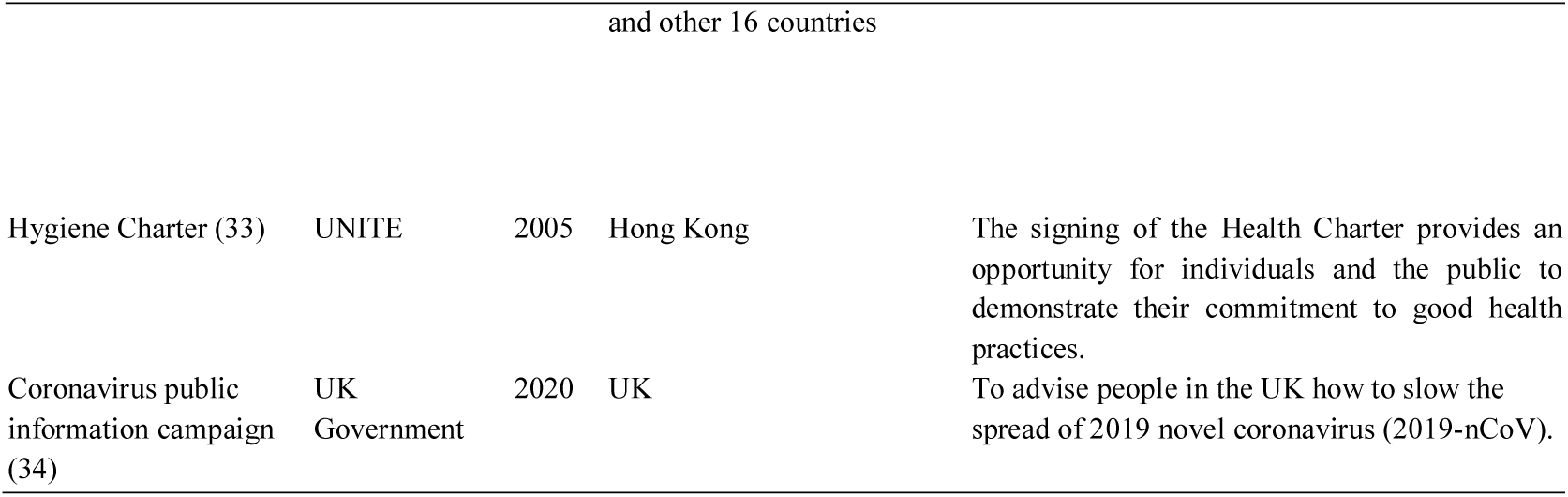
Health education and health promotion programs launched after COVID-19 and SARS.

Because of the limitations in the type of study we could not perform the assessment of quality of evidence using GRADE. Overall, the included surveys had acceptable design and reporting quality (*Table 3*), but the sampling methods were not described with sufficient details in some studies. Many online surveys had a large sample size but their reliability was questioned.

**Table 3.** Risk of bias in the included studies.

| Study | Item 1 | Item 2 | Item 3 | Item 4 | Item 5 | Item 6 | Item 7 | Item 8 | Item 9 | Item 10 |
| --- | --- | --- | --- | --- | --- | --- | --- | --- | --- | --- |
| Chen 2020(9) | Yes | No | Yes | Yes | Yes | Yes | Yes | Yes | Yes | Yes |
| Zhong 2020(10) | Yes | No | No | Yes | UN | Yes | UN | Yes | Yes | Yes |
| Zheng 2020(11) | No | No | Yes | Yes | No | Yes | No | Yes | Yes | UN |
| Qi 2020(12) | Yes | No | Yes | Yes | No | No | No | Yes | Yes | UN |
| Zhu 2020(13) | No | No | Yes | Yes | No | Yes | No | Yes | Yes | UN |
| Yang 2020(14) | No | No | Yes | Yes | No | Yes | No | Yes | Yes | UN |
| Song 2003(15) | Yes | Yes | Yes | Yes | Yes | No | Yes | Yes | No | No |
| Zhang 2004(16) | Yes | Yes | Yes | Yes | Yes | Yes | Yes | Yes | No | No |
| Chan 2007(17) | Yes | No | Yes | Yes | UN | Yes | Yes | Yes | Yes | — |
| Du 2009(18) | Yes | Yes | Yes | Yes | Yes | Yes | Yes | Yes | — | — |
| Gautret 2013(19) | Yes | UN | Yes | Yes | Yes | — | Yes | Yes | — | Yes |
| Bener 2004(20) | Yes | Yes | Yes | Yes | Yes | Yes | Yes | Yes | Yes | Yes |
| Sahin 2015(21) | Yes | UN | Yes | Yes | Yes | — | Yes | Yes | — | Yes |
| Jardine 2015(22) | Yes | Yes | Yes | Yes | — | — | Yes | Yes | UN | — |
| Alqahtani 2016(23) | Yes | UN | Yes | Yes | UN | — | Yes | Yes | Yes | — |
| Huang 2016(24) | Yes | No | Yes | Yes | No | No | Yes | Yes | Yes | — |
| Alhomoud 2017(25) | Yes | No | No | Yes | — | — | Yes | Yes | UN | — |
| Bawazir 2018(26) | Yes | No | Yes | Yes | — | Yes | Yes | Yes | Yes | — |
| Migault 2019(27) | Yes | No | No | Yes | UN | — | Yes | — | — | — |
| Zhu 2004(28) | Yes | No | Yes | Yes | No | No | Yes | Yes | Yes | — |
| Zhong 2004(29) | Yes | No | Yes | Yes | No | Yes | Yes | Yes | No | No |
| Alqahtani 2017(30) | Yes | Yes | Yes | UN | UN | Yes | UN | Yes | Yes | Yes |
| Almutairi 2015(31) | UN | Yes | Yes | Yes | UN | Yes | UN | Yes | Yes | Yes |
| Al-Mohrej 2016(32) | Yes | Yes | Yes | Yes | UN | Yes | UN | Yes | Yes | Yes |

Note: UN: Unknown; —: Not applicable; 1. Sample representativeness; 2. Sampling method; 3. Sample size; 4. Study subjects and setting; 5. Coverage of the identified sample; 6. Using of standardized criteria for the measurement; 7. Reliability of the measurement; 8. Appropriateness of
the statistical analysis; 9. Confounding factors/subgroups/differences; 10. Objectiveness of the criteria of subpopulations

## Discussion and Action Plan

### Boosting health education to improve public health awareness for infectious diseases

Improved public health awareness for emerging infectious diseases plays a critical role in at least two ways. On one hand, appropriate behavior of the public, like adhering to self-quarantine and practicing the necessary hygienic habits will definitely slow down the spreading and help control the epidemic. On the other hand,outbreaks of emerging infectious diseases can often lead to negative social phenomena such as fear, stigma, and discrimination (36). Individuals who are feared and stigmatized may deny early clinical symptoms, delay seeking care, and remain in the community undetected, which may aggravate the spread of infection. Better knowledge and awareness promote understanding and communication between different groups of people and improve mental health. WHO had developed guidelines for preventing and addressing social stigma (45), and governments, media, and local organizations working on the SARS-CoV-2 epidemic should follow the recommendations. To control the spreading of COVID-19, boosting public health education should be a priority.

### Health education needed for special groups

The general public may lack adequate knowledge about emerging infectious diseases, and this situation may be even worse in special groups, like children, the elderly, travelers, and other vulnerable groups. This review found that the uneducated and young people lacked appropriate information about infectious diseases and adequately behavior in this outbreak (26). Children may have difficulties in understanding of mass health education. Parents have an obligation to explain disease prevention measures to their children. Furthermore, they should help children to be aware of infectious diseases. We identified three studies (38-40) that evaluated the health care staff’s knowledge of SARS or MERS, and found out the importance of providing related health education to them.

In the context of globalization, the speed and mode of infectious disease transmission have changed substantially. An outbreak of an infectious disease commonly restricted in epidemic focus, and travel bureaus or visa agencies usually post an alert or warning when the destination undergoes an epidemic, reminding people to be cautious or change their travel plans. The survey of MERS knowledge among pilgrimage travelers could insinuate a similar condition in SARS-CoV-2. SARS-CoV-2 is highly contagious, and essentially everyone is susceptible as there is no vaccination or natural immunity. Therefore, travelers should pay attention to the prevalence of SARS-CoV-2 in tourist destinations when arranging family travel or overseas study tours, especially while the SARS-CoV-2 epidemic is still ongoing.

### Improving access to more reliable information

Two studies (43,44) showed that health information from nonprofit, government and academic websites was more accurate than privately owned commercial websites and media websites. With the development of new media, people can now draw information from multiple sources, and through mobile devices basically at any time and place. Social media like Facebook, Twitter and WeChat are full of SARS-CoV-2 news and health education resources, which is convenient for the phone users. However, fake news and gossips are spreading fast, just like the virus. We appeal for the media and publishers to take deep consideration before post any information to the public. Official websites of the WHO and the national CDC are updating information about the epidemic and necessary preventive measures, and it is suggested for the publisher to cite and follow them.

In the event of a public health emergency, professional agencies, government departments, and authoritative media should fully cooperate according to the information needed by the public at different stages of the crisis under the unified coordination of the government. This way the core information required by the public will transmit to the public in a timely, accurate, and appropriate manner (18).

### Launching health education campaigns

Although we have collected data on some health education and promotion campaigns or programs in the world, we still need a wide influenced and persistent-actioning campaign to face the outbreak of the novel infectious disease. The outbreaks of SARS, MERS, Ebola and COVID-19 have given us every time a lesson for solidarity and collaboration. Suffering from the pain of lost lives, health education campaigns should be launched as early as possible to slow down the spread immediately.

## Conclusions

Our rapid review provides evidence of the importance and urgency of health education. Boosting health education to improve public health awareness for infectious diseases is urgently needed, especially for some vulnerable groups. Considering that information from the social media may be unreliable, the public including parents should obtain information from the official websites of authorities such as the World Health Organization and national Center for Disease Control, or from other sources endorsed by these authorities, rather than from a general search of the internet or social media. For educating and cultivating children, parents need to educate their children on the importance of evidence-based information on COVID-19 and help them practice preventive measures and hygiene behaviors.

## Data Availability

This study is a rapid review based on the original researches. All the data are real and are extracted from existing original researches.

## Author contributions

(I) Conception and design: Zhengxiu Luo, Yaolong Chen and Enmei Liu; (II) Administrative support: Enmei Liu; (III) Systematic Searching: Weiguo Li, Jing Liao, Qinyuan Li; (IV) Collection and assembly of data: Jing Liao and Weiguo Li; (V) Data analysis and interpretation: Weiguo Li, Jing Liao and Muna Baskota; (VI) Manuscript writing: All authors; (VII) Final approval of manuscript: All authors.

## Acknowledgments

We thank Janne Estill, from Institute of Global Health of University of Geneva, for providing guidance and comments for our review. We thank all the authors for their wonderful collaboration.

## Footnote

### Ethical Statement

The authors are accountable for all aspects of the work in ensuring that questions related to the accuracy or integrity of any part of the work are appropriately investigated and resolved.

## Supplementary Material 1-Search strategy

### PubMed

#1 “COVID-19”[Supplementary Concept]

#2 “Severe Acute Respiratory Syndrome Coronavirus 2”[Supplementary Concept]

#3 “Middle East Respiratory Syndrome Coronavirus”[Mesh]

#4 “Severe Acute Respiratory Syndrome”[Mesh]

#5 “SARS Virus”[Mesh]

#6 “COVID-19”[Title/Abstract]

#7 “SARS-COV-2”[Title/Abstract]

#8 “Novel coronavirus” [Title/Abstract]

#9 “2019-novel coronavirus” [Title/Abstract]

#10 “coronavirus disease-19” [Title/Abstract]

#11 “coronavirus disease 2019” [Title/Abstract]

#12 “COVID 19” [Title/Abstract]

#13 “Novel CoV” [Title/Abstract]

#14 “2019-nCoV” [Title/Abstract]

#15 “2019-CoV” [Title/Abstract]

#16 “Wuhan-Cov” [Title/Abstract]

#17 “Wuhan Coronavirus” [Title/Abstract]

#18 “Wuhan seafood market pneumonia virus” [Title/Abstract]

#19 “Middle East Respiratory Syndrome” [Title/Abstract]

#20 “MERS” [Title/Abstract]

#21 “MERS-CoV” [Title/Abstract]

#22 “Severe Acute Respiratory Syndrome” [Title/Abstract]

#23 “SARS” [Title/Abstract]

#24 “SARS-CoV” [Title/Abstract]

#25 “SARS-Related” [Title/Abstract]

#26 “SARS-Associated” [Title/Abstract]

#27 #1-#26/ OR

#28 “Health education” [Title/Abstract]

#29 “Health promotion” [Title/Abstract]

#30 “Parent education” [Title/Abstract]

#31 “Patient education” [Title/Abstract]

#32 “Health education” [Mesh]

#33 “Health promotion” [Mesh]

#34 #28-#33/ OR

#35 #27 AND #34

### EMBASE

#1 ‘middle east respiratory syndrome coronavirus’/exp

#2 ‘severe acute respiratory syndrome’/exp

#3 ‘sars coronavirus’/exp

#4 ‘COVID-19’:ab,ti

#5 ‘SARS-COV-2’:ab,ti

#6 ‘novel coronavirus’:ab,ti

#7 ‘2019-novel coronavirus’:ab,ti

#8 ‘coronavirus disease-19’:ab,ti

#9 ‘coronavirus disease 2019’:ab,ti

#10 ‘COVID19’:ab,ti

#11 ‘novel cov’:ab,ti

#12 ‘2019-ncov’:ab,ti

#13 ‘2019-cov’:ab,ti

#14 ‘wuhan-cov’:ab,ti

#15 ‘wuhan coronavirus’:ab,ti

#16 ‘wuhan seafood market pneumonia virus’:ab,ti

#17 ‘middle east respiratory syndrome’:ab,ti

#18 ‘middle east respiratory syndrome coronavirus’:ab,ti

#19 ‘mers’:ab,ti

#20 ‘mers-cov’:ab,ti

#21 ‘severe acute respiratory syndrome’:ab,ti

#22 ‘sars’:ab,ti

#23 ‘sars-cov’:ab,ti

#24 ‘sars-related’:ab,ti

#25 ‘sars-associated’:ab,ti

#26 #1-#25/ OR

#27 ‘health promotion’/exp

#28 ‘health education’:ab,ti

#29 ‘health promotion’:ab,ti

#30 ‘parent education’:ab,ti

#31 ‘patient education’:ab,ti

#32 #27-#31/ OR

#33 #26 AND #32

### Web of Science

#1 TOPIC: “COVID-19”

#2 TOPIC: “SARS-COV-2”

#3 TOPIC: “Novel coronavirus”

#4 TOPIC: “2019-novel coronavirus”

#5 TOPIC: “coronavirus disease-19” [Title/Abstract]

#6 TOPIC: “coronavirus disease 2019” [Title/Abstract]

#7 TOPIC: “COVID19” [Title/Abstract]

#8 TOPIC: “Novel CoV”

#9 TOPIC: “2019-nCoV”

#10 TOPIC: “2019-CoV”

#11 TOPIC: “Wuhan-Cov”

#12 TOPIC: “Wuhan Coronavirus”

#13 TOPIC: “Wuhan seafood market pneumonia virus”

#14 TOPIC: “Middle East Respiratory Syndrome”

#15 TOPIC: “MERS”

#16 TOPIC: “MERS-CoV”

#17 TOPIC: “Severe Acute Respiratory Syndrome”

#18 TOPIC: “SARS”

#19 TOPIC: “SARS-CoV”

#20 TOPIC: “SARS-Related”

#21 TOPIC: “SARS-Associated”

#22 #1-#21/ OR

#23 TOPIC: “Health education”

#24 TOPIC: “Health promotion”

#25 TOPIC: “Parent education”

#26 TOPIC: “Patient education”

#27 #23-#26/ OR

#28 #22 AND #27

### Cochrane

#1 MeSH descriptor: [Middle East Respiratory Syndrome Coronavirus] explode all trees

#2 MeSH descriptor: [Severe Acute Respiratory Syndrome] explode all trees

#3 MeSH descriptor: [SARS Virus] explode all trees

#4 “COVID-19”:ti,ab,kw

#5 “SARS-COV-2”:ti,ab,kw

#6 “Novel coronavirus”:ti,ab,kw

#7 “2019-novel coronavirus” :ti,ab,kw

#8 “Novel CoV” :ti,ab,kw

#9 “2019-nCoV” :ti,ab,kw

#10 “2019-CoV” :ti,ab,kw

#11 “coronavirus disease-19” :ti,ab,kw

#12 “coronavirus disease 2019” :ti,ab,kw

#13 “COVID19” :ti,ab,kw

#14 “Wuhan-Cov” :ti,ab,kw

#15 “Wuhan Coronavirus” :ti,ab,kw

#16 “Wuhan seafood market pneumonia virus” :ti,ab,kw

#17 “Middle East Respiratory Syndrome” :ti,ab,kw

#18 “MERS”:ti,ab,kw

#19 “MERS-CoV”:ti,ab,kw

#20 “Severe Acute Respiratory Syndrome”:ti,ab,kw

#21 “SARS” :ti,ab,kw

#22 “SARS-CoV” :ti,ab,kw

#23 “SARS-Related”:ti,ab,kw

#24 “SARS-Associated”:ti,ab,kw

#25 #1-#24/ OR

#26 “Health education”:ti,ab,kw

#27 “Health promotion”:ti,b,kw

#28 “Parent education”:ti,ab,kw

#29 “Patient education”:ti,ab,kw

#30 MeSH descriptor: [Health promotion] explode all trees

#31 #26-#30/ OR

#32 #25 AND #31

### CBM

#1 “新型冠状病毒”[常用字段:智能]

#2 “COVID-19”[常用字段:智能]

#3 “COVID 19”[常用字段:智能]

#4 “2019-nCoV”[常用字段:智能]

#5 “2019-CoV”[常用字段:智能]

#6 “SARS-CoV-2”[常用字段:智能]

#7 “武汉冠状病毒”[常用字段:智能]

#8 “中东呼吸综合征冠状病毒”[不加权:扩展]

#9 “中东呼吸综合征”[常用字段:智能]

#10 “MERS”[常用字段:智能]

#11 “MERS-CoV”[常用字段:智能]

#12 “严重急性呼吸综合征”[不加权:扩展]

#13 “SARS 病毒”[不加权:扩展]

#14 “严重急性呼吸综合征”[常用字段:智能]

#15 “SARS”[常用字段:智能]

#16 #1-#15/ OR

#17 “健康教育”[不加权:扩展]

#18 “健康教育”[常用字段:智能]

#19 “健康促进”[常用字段:智能]

#20 “卫生教育”[常用字段:智能]

#21 “家长教育”[常用字段:智能]

#22 “患者教育”[常用字段:智能]

#23 “科普知识”[常用字段:智能]

#24 “科普教育”[常用字段:智能]

#25 “科普宣传”[常用字段:智能]

#26 #17-#25/ OR

#27 #16 AND #26

### WanFang

#1 “新型冠状病毒”[主题]

#2 “COVID-19”[主题]

#3 “COVID-19”[主题]

#4 “2019-nCoV”[主题]

#5 “2019-CoV”[主题]

#6 “SARS-CoV-2”[主题]

#7 “武汉冠状病毒”[主题]

#8 “中东呼吸综合征”[主题]

#9 “MERS”[主题]

#10 “MERS-CoV”[主题]

#11 “严重急性呼吸综合征”[主题]

#12 “SARS”[主题]

#13 #1-#12/ OR

#14 “健康教育”[主题]

#15 “健康促进”[主题]

#16 “卫生教育”[主题]

#17 “家长教育”[主题]

#18 “患者教育”[主题]

#19 “科普教育”[主题]

#20 “科普知识”[主题]

#21 “科普宣传”[主题]

#22 #14-#21/ OR

#23 #13 AND #22

### CNKI

#1 “新型冠状病毒”[主题]

#2 “COVID-19”[主题]

#3 “COVID-19”[主题]

#4 “2019-nCoV”[主题]

#5 “2019-CoV”[主题]

#6 “SARS-CoV-2”[主题]

#7 “武汉冠状病毒”[主题]

#8 “中东呼吸综合征”[主题]

#9 “MERS”[主题]

#10 “MERS-CoV”[主题]

#11 “严重急性呼吸综合征”[主题]

#12 “SARS”[主题]

#13 #1-#12/ OR

#14 “健康教育”[主题]

#15 “健康促进”[主题]

#16 “卫生教育”[主题]

#17 “家长教育”[主题]

#18 “患者教育”[主题]

#19 “科普教育”[主题]

#20 “科普知识”[主题]

#21 “科普宣传”[主题]

#22 #14-#21/ OR

#23 #13 AND #22

